# Wastewater Tiling Amplicon Sequencing Reveals Longitudinal Dynamics of SARS-CoV-2 Variants Prevalence in the Community

**DOI:** 10.1101/2023.10.30.23297759

**Authors:** Yu Wang, Gaofeng Ni, Wei Tian, Haofei Wang, Jiaying Li, Phong Thai, Phil M. Choi, Greg Jackson, Shihu Hu, Bicheng Yang, Jianhua Guo

## Abstract

SARS-CoV-2 continues to evolve, while the decline in clinical sequencing efforts hampers public health sectors to prepare for the menace of ongoing variant emergence and future COVID-19 surges.^1^ Wastewater-based epidemiology (WBE) has been proposed to provide complementary insights on the variants being transmitted in communities.^2^ However, limited research has been dedicated to the use sequencing methods for tracking disease prevalence and variant dynamics in wastewater, particularly on a large scale. Here, we employed a tiling amplicon sequencing to track the dynamics of variant of concern (VOC) in wastewater collected from Queensland, Australia from 2020 to 2022. RNA concentrations in wastewater measured by ATOPlex showed a stronger correlation and greater consistency with the number of daily new cases than a PCR-based method. The VOC dynamics observed in wastewater were largely in line with clinical reports. These findings support that WBE and sensitive sequencing methods can serve as a long-term approach for disease surveillance, thus aiding in disease outbreak prevention, control, and management.

## 1. Introduction

The World Health Organization (WHO) designates evolved variants with genetic mutations associated with increased transmissibility, pathogenicity and immune escape as variants of concern (VOCs). Since the appearance of the first VOC, namely Alpha variant, in 2020, emergent variants have caused several global surges in COVID-19.^3^ The monitoring of VOCs and related genetic characteristics typically depends on extensive individual clinical sequencing.^4^ However, clinical surveillance has an unavoidable risk of sampling bias and is becoming less effective in revealing circulating variants, as the global response to COVID-19 strategies are adjusted.^5,6^ This situation is even worse where medical resources are scarce, as large-scale clinical sequencing is laborious, expensive and time-consuming.^4,6^ The risk of new variants to emerge calls for sustained monitoring strategies to inform timely preventive measures.^7^

Wastewater-based epidemiology (WBE) enables effective identification and quantification of disease prevalence at a community level.^8^ It has been implemented as a surveillance program worldwide, such as in the USA, Australia, and Europe in response to COVID-19.^9–12^ The use of PCR-based methods has facilitated WBE in monitoring the disease burden in communities and providing early warning of disease outbreaks before clinical surveillance.^13–15^ However, mutations or degradation of the N gene, the widely-used PCR primer binding region, will reduce the primer binding efficiency. This may lead to false negative PCR results and impeding the timeliness of WBE.^16,17^ Moreover, despite advancements in variant-specific PCR assays for identifying different VOCs in wastewater, determining transmission links and timely detecting emerging variants in a community remain significant challenges.^18,19^

In contrast, sequencing technologies can directly recover viral genomes from wastewater, allowing the discerning of different variants and their co-occurrence.^5^ The SARS-CoV-2 RNA in wastewater is typically present in low concentration and is readily fragmented, which requires a sensitive sequencing method for the identification of SARS-CoV-2 and the recovery of a high-quality genome.^20^. Tiling amplicon sequencing, such as ARTIC, ATOPlex and Midnight, have been proposed as sensitive and cost-efficient approaches for recovering viral genomes from wastewater.^21–23^ By using amplicon sequencing technologies, wastewater surveillance offers valuable information on disease prevalence and can potentially inform transmission links in communities. For example, research has demonstrated a strong correlation between the proportion of wastewater samples containing specific variant signals and the corresponding percentage observed in clinical samples.^24,25^ The recovery of high-coverage genomes from wastewater uncovers various VOCs introduced across different regions.^26,27^ Building-level wastewater sequencing facilitates early detection of infection events, enabling prompt actions to identify positive cases through clinical testing, then relocate infected individuals, potentially preventing potential outbreaks.^5^ Furthermore, WBE surveillance within a university campus has been shown to track the temporal profile of different variants in a defined region through intensive sampling.^5,14^ However, still limited effort has been made to assess the application of wastewater sequencing in tracking variant dynamics and comparing it with clinical sequencing data. More importantly, it remains unclear whether collecting wastewater from the strategically chosen sampling sites can effectively track the variant dynamics in large-scale areas over the long term.

This study conducted a longitudinal investigation, collecting wastewater samples over a span of three years (from mid-2020 to late-2022), from three municipal sites in Queensland, Australia. The objective of this study was to evaluate the effectiveness of long-term wastewater sequencing surveillance at sentinel sites in estimating disease prevalence and monitoring the dynamics of SARS-CoV-2 variants at the state level. By using ATOPlex sequencing, we successfully tracked the circulating dynamics of SARS-CoV-2 variants and have the potential to detect emerging variants at an early stage in wastewater samples. Furthermore, wastewater sequencing exhibited stronger correlation and consistency in estimating disease prevalence trends compared to a PCR-based method in different sampling sites. Overall, this study offers valuable insights into the use of WBE for monitoring disease dynamics in the community, particularly in the context of easing public health measures.

## 2. Methods

### 2.1 Study area and sample collection

Wastewater samples were collected from six wastewater treatment plants (WWTPs) located in three municipalities within Queensland, Australia (Figure S1). A total of 113 samples that tested positive for SARS-CoV-2 via RT-qPCR testing were obtained, including WWTPs A (n=11), B (n=3), C (n=40), D (n=4), E (n=40), and F (n=15), respectively (Figure 1). Samples from WWTPs C and E were collected between July 2020 and September 2022, while WWTPs A, B and D ceased collection in 2022. To further investigate the presence of emerging variants that have not been locally endemic in Queensland, 15 grab samples were collected in November-December 2022 from the largest WWTP in Queensland, WWTP F, which serves the busiest locations of Queensland, including the capital city Brisbane (BNE) and an international airport (Figure 1). The sampling sites and the served population of each WWTP included in the study are depicted in Figure S1. In 2020 and for the majority of 2021, Queensland implemented stringent public health measures, such as tight border restrictions, during which 14 days of quarantine was required to enter the state. Consequently, the collected samples had a low RNA concentration. Moreover, the RNA in wastewater was subject to degradation during storage, which further contributed to negative RT-qPCR results at the time of sequencing in September 2022.^12^ Therefore, only a few samples from 2020/2021 were included to test the sensitivity of ATOPlex sequencing.

**Figure 1.**
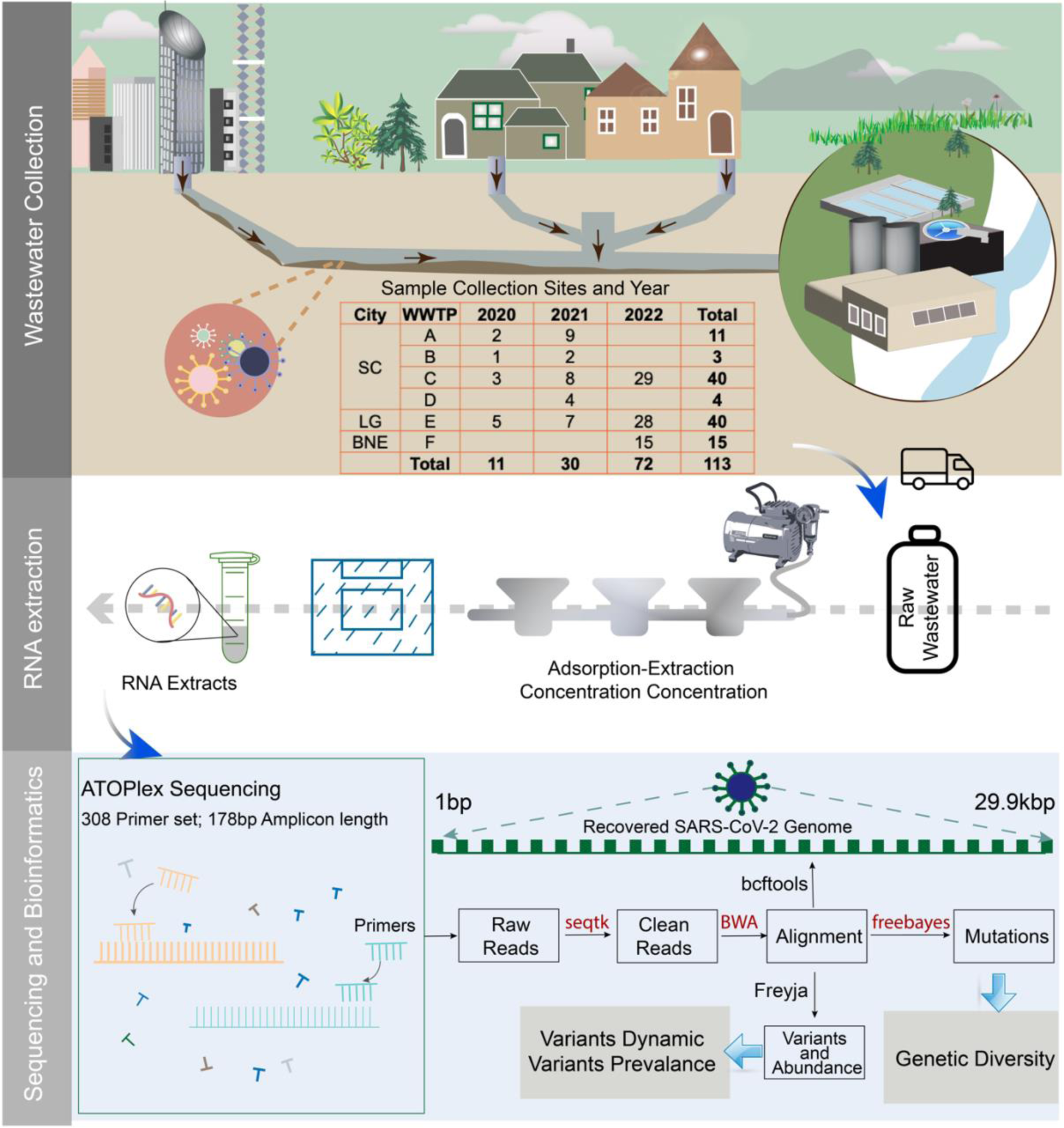
Overview of the wastewater surveillance program. Top panel: Concept of the WBE and collection sites of raw wastewater samples. Middle panel: Wastewater concentration and RNA extraction. Bottom panel: ATOPlex sequencing and bioinformatics analysis.

### 2.2 Wastewater sample preparation, concentration, and RNA extraction

Samples were transported on ice and stored at -20 °C until further study. The frozen samples were first thawed at 4 °C followed by wastewater concentration and RNA extraction as described in previous studies (Figure 1).^28,29^ Briefly, 0.5 mL of 25 mM dissolved MgCl2 was added to the aliquots of 50 mL raw wastewater samples and incubated at room temperature for a few minutes. Samples were then filtered through a 0.45-µm pore-size, 47-mm diameter electronegative HA membrane (HAWP04700; Merck Millipore Ltd, Sydney, Australia) using a multichannel filter funnel. All equipment was autoclaved at 121 °C for 20 minutes prior to use. The membrane was removed from the filtration apparatus, rolled, shredded and placed into a 2 ml beat tube from RNeasy PowerMicrobiome Kit (Qiagen) using two sterile tweezers. RNA was then extracted following the manufacturer’s instructions with the following modifications:

(1) 100 μL of phenol–chloroform–isoamyl alcohol (25:24:1) was added before the lysis step to protect RNA integrity; (2) Beat-beat for 60s with Minilys® homogenizer at medium speed; (3) IRS solution was increased to 200 μL to remove more inhibitors, and (4) RNA eluted with 50 μL Nuclease free water (Biolab) and pass-through spin column twice to ensure maximum RNA yield. RNA extracts were stored at -80 °C before sequencing.

### 2.3 Positive control preparation

A commercially available RNA positive control was used to prepare a serial dilution with 5 orders of gradient, as shown in Figure S1 (b). Briefly, 10 *μ*L RNA was reverse transcribed to cDNA using the high-capacity cDNA reverse transcription kit (Catalog No. 4368814, Applied Biosystems™) and serially diluted by 5 orders of magnitude with nuclease-free water.^26^ The concentration of each positive control gradient was measured by droplet digital Polymerase Chain Reaction (dd-PCR) assay, as described in a previous study,^28^ with the concentration ranging from 6235.58 to 0.63 copies/*μ*L.

### 2.4 ATOPlex sequencing

A total of 113 wastewater RNA extracts and 5 positive control gradients (in triplicate) were sequenced using the ATOPlex SARS-CoV-2 full-length genome panel (MGI, China) as described previously. Briefly, sample RNA was reverse transcribed to cDNA with random hexamers (5’-NNNNNN-3’). Then, 200 copies of Lambda phage DNA were dosed into each sample as an internal control to set a baseline for result quantification. The resulting product undergoes two steps of PCR amplification followed by purification, with the first PCR of 13 cycles for target enrichment, and the second PCR of 27 cycles for barcoding and product amplification. The process was completed using the automation sample processing system MGISP-960. The purified final PCR products were quantified using Qubit Quant-iT™ dsDNA Assay Kits (ThermoFisher, Q33120). Subsequently, PCR products of 96 samples were pooled into two pools, with each pool containing 48 samples and 400 ng of library. The pooled dsDNA libraries were denatured to generate linear ssDNA, circularized using a splint oligo that is complementary to the start and end of linear ssDNA, followed by digestion reaction and purification to remove non-circularized DNA debris. The process was completed manually using MGIEasy Dual Barcode Circularization Kit (MGI, China) to produce purified single-strand circularized products. The ssDNA circularized product was then used as a template to generate DNA nanoball (DNBs) by performing a rolling circle amplification reaction. The DNBs were introduced into flow cell containing patterned arrays of DNB binding sites for sequencing. The sequencing was done on a DNBSeq-G400 sequencer running for 24 hours to get 8M reads for each sample at the MGI Australia Demo Lab.

### 2.5 Data analysis and interpretation

As part of the surveillance program, wastewater monitoring of SARS-CoV-2 using the RT-qPCR method was applied, and the result was obtained from Queensland Public Health and Scientific Services. The sequencing raw data were analysed with the bioinformatic pipeline for multiplex-PCR data developed by MGI (https://github.com/MGI-tech-bioinformatics/SARS-CoV-2_Multi-PCR_v1.0). The established pipeline is used to analyse raw sequencing data, including genome alignment, mutation calling, consensus genome generation and identification (Figure 1).^28,30^ Viral concentration in wastewater was calculated based on normalized SARS-CoV-2 sequencing reads. The higher coverage and depth underpin a greater likelihood of accurate sequencing and variant detection at that position.^25^ To ensure data precision, only samples identified as positive and genomes with coverage (10X) of at least 20% were included for variant identification and investigation of genetic diversity. Freyja (V1.3.11), a well-established pipeline bioinformatics tool, was employed to determine co-circulating variants and corresponding prevalence in wastewater samples.^5^ COVID-19 case data in the study areas were sourced from the Queensland Government Open Data Portal (https://www.data.qld.gov.au). Clinical genomic data from Queensland were sourced from the GISAID database.^31^ The clinical genomes were then grouped by collection date, and mutations in the groups were obtained using snp-sites (V2.5.1),^32^ the mutation frequency was calculated using a custom script. Genetic diversity of the wastewater samples and clinical samples was measured by the richness (count of the number of mutations present in the sample) and Shannon Entropy (H(x) = - Σ p(x) log₂(p(x), where p(x) is the allele frequency at the position x), the distribution of each mutation and degree of genetic diversity in the sample).

### 2.6 Data availability

All the raw sequencing data are available at the National Center for Biotechnology Information (NCBI) Sequence Read Archive (SRA) under BioProject number PRJNA945486.

## 3. Results

### 3.1 Accurate viral RNA concentration estimation and genome recovery

To verify whether the chosen sequencing method can accurately quantify the virus concentration, a linear regression analysis is used to predict the relationship between the RNA concentration measured by dd-PCR with the sequencing reads. The result revealed a linear regression correlation (R^2^adj=0.99) between the normalized SARS-CoV-2 sequencing reads and absolute RNA concentration measured by dd-PCR from concentration gradients D1 (6235.58 cp/*μ*L) to D5 (0.63 cp/*μ*L), as shown in Figure 2-a. The result is consistent with our past study and further verifies the stability of the ATOPlex sequencing method.^28^ Furthermore, ATOPlex sequencing can accurately recover the SARS-CoV-2 genome (median 10X coverage of 98.19%) even with mutations in comparison to the reference genome (MN908947.3) and at a concentration as low as 0.63 cp/*μ*L (Figure 2-b). The accuracy estimation of viral concentration and derived genomes facilitated the subsequent analysis for qualitative and quantitative analysis of the SARS-CoV-2 virus.

**Figure 2.**
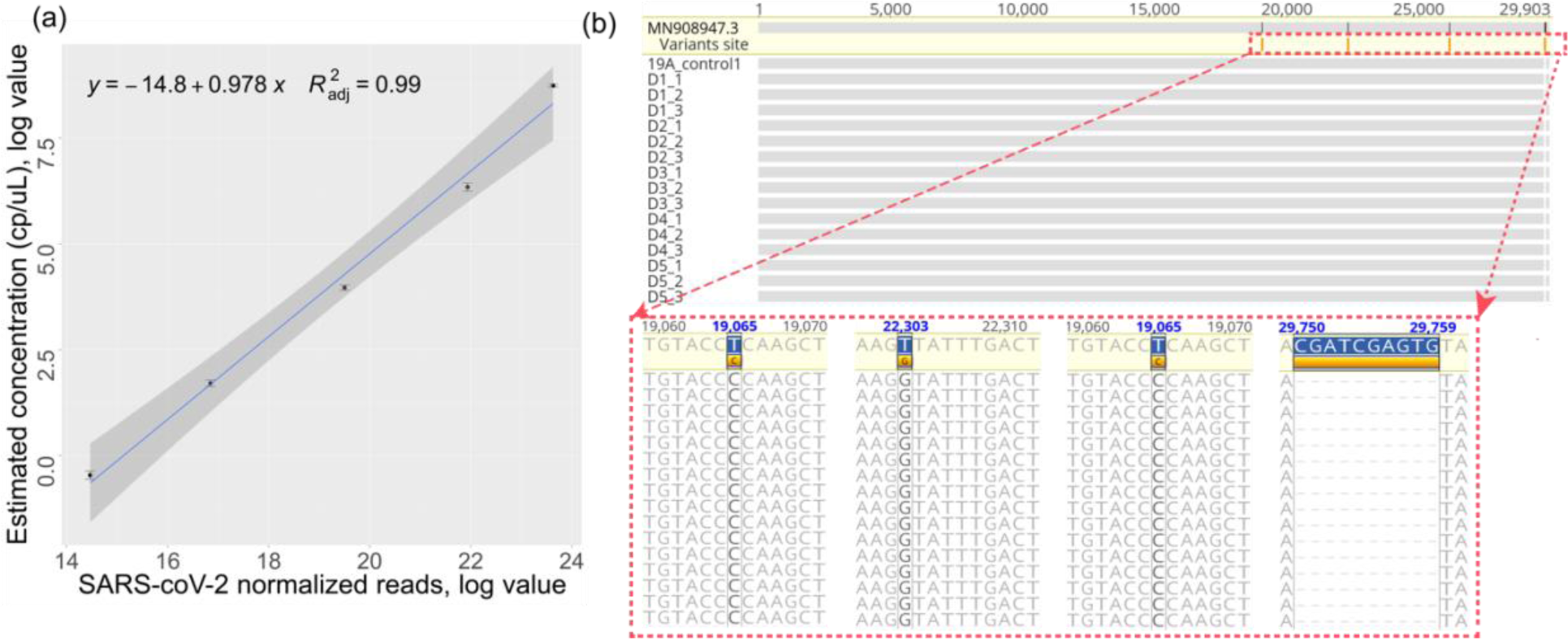
(a) Linear relationship between normalized ATOPlex SARS-CoV-2 reads and viral concentration measured by dd-PCR at log value. (b) Sequence alignments of the reference genome (MN908947.3) and ATOPlex recovered genome across serial dilution D1-D5 align to the positive control 19A genome. Nucleotides that differ from the positive control genome are highlighted in black and have detailed alignment information enlarged.

### 3.2 Identification of the SARS-CoV-2 in wastewater samples

ATOPlex classify samples as positive, negative, or indeterminate by normalizing the number of SARS-CoV-2 reads to the spike-in lambda phage DNA control reads and assessing genome coverage.^28,30^ In summary, out of the 113 wastewater samples 109 tested positive, while only 4 samples were categorized as indeterminate due to low genome coverage. Wherein some samples might have become highly degraded and fragmented during long-time storage. To gain further insights into the completeness of the recovered viral genome, the genome coverage was visualized (Figure S2). Median SARS-CoV-2 genome coverage (10X) was 52.66% (range 0.69–99.88%). Figure 3 illustrates three examples of the distribution of sequencing reads with a different genome coverage (10X). It is apparent that ATOPlex still can identify the SARS-CoV-2 virus even in instances where the primer regions N1 and N2 commonly used in PCR-based methods were degraded (Figure 3). ATOPlex sequencing can provide high-quality genomes, except in several cases where the genome region has undergone significant degradation.

**Figure 3.**
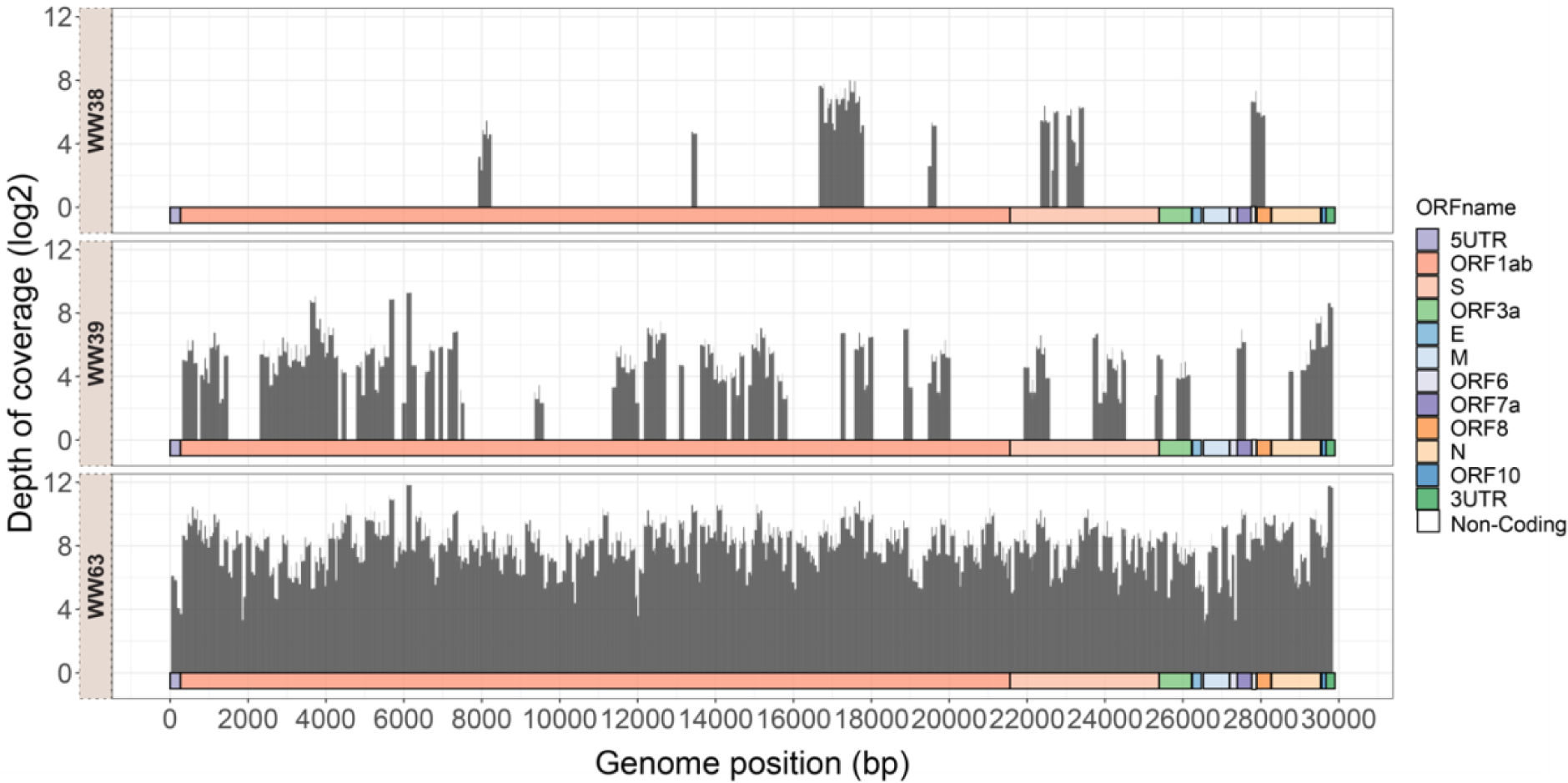
Example of genome coverage (10X) of the wastewater samples. The 10X genome coverage for WW38, WW39 and WW63 are 14.63%, 50.62 and 99.88% respectively. The x-axis represents the coverage of every single nucleotide position with respect to the reference genome (MN908947.3) and the y-axis represents the sequencing depth at the log2 value at each position.

### 3.3 Wastewater sequencing reveals the viral load and prevalence trend

To assess the consistency of the disease prevalence between wastewater surveillance and clinical surveillance, we compared RNA concentration estimated by wastewater sequencing with clinical daily new case numbers. In 2020 and 2021, Queensland was marked by strict disease control measures, such as border closures, gathering restrictions, and targeted mandatory vaccinations, thus minimising local transmission (Table S1). However, following the opening of the Queensland border in December 2021, there is a dramatic increase in weekly total cases, with a nearly 150% increase observed in the week after the reopening compared to the week prior (Figure 4-a). Concurrently, a consistent upward trend in RNA concentration was observed in wastewater samples (Figure 4-a). In addition, COVID-19 exhibited four main surges during the study period in terms of both clinical and wastewater surveillance, which is thought to be associated with the emergence of new variants (Figure 4-a). For example, BA.1 was first detected in clinical testing in December 2021 and rapidly spread after that. Along with low levels of other variants, BA.1 was observed in all areas inspected in this study, resulting in the first COVID-19 epidemic wave in Queensland (Figure 4-a, Figure S3). Interestingly, the RNA concentration in wastewater samples in February 2022 exceeded the clinical case line on a monthly scale (Figure 4-b, c), indicating a higher estimated prevalence of COVID-19 through wastewater surveillance than clinically confirmed cases.

**Figure 4.**
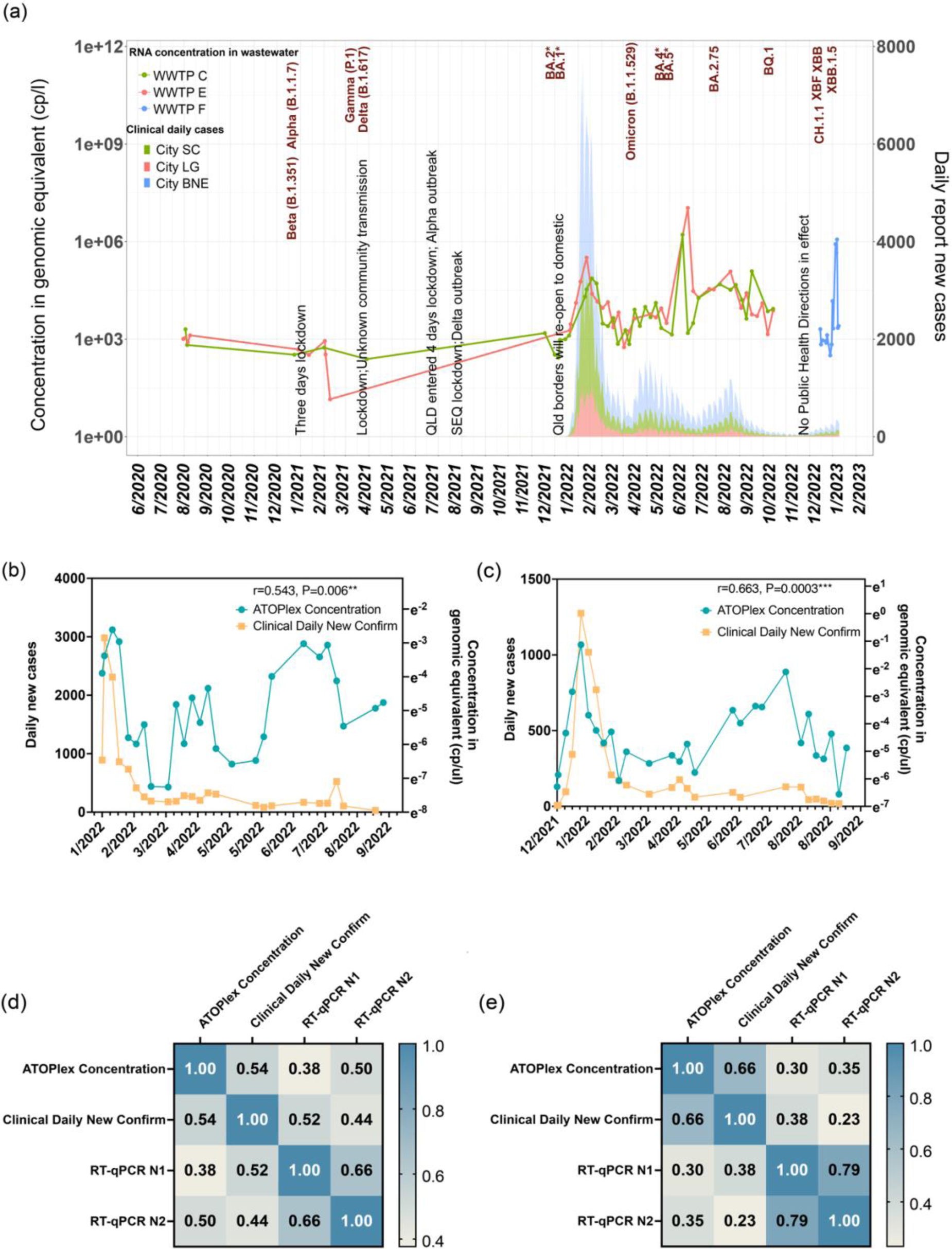
Epidemiological curves depicting disease prevalence in the study regions. (a) The line graph represents the wastewater epidemiological curve and the area plot shows the clinical daily new confirmed cases. The major public health measures (shown in black), as well as the first detection date of each emerging variant (shown in red) identified through clinical sequencing during the study period are labelled. (b) and (c) zoom in on the curve measured by wastewater sequencing (in blue) and clinical tests (in yellow) in SC and LG regions, respectively. (d) and (e) shows The Pearson correlation coefficient (|r|) matrix for each measuring method with the daily new cases in SC and LG.

Moreover, in terms of correlation with different clinical epidemiology factors, WBE has a higher performance in estimating daily new cases compared to active or cumulative clinical cases.^33^ To evaluate the correlation between RT-qPCR and ATOPlex in estimating the the new cases reported each day, Pearson correlation coefficient analysis was employed. The sequencing method exhibited a higher correlation with the daily new cases (Figure 4-d, e), with a |r| equal to 0.54 (p=0.006) and 0.663 (p=0.0003)) in the SC and LG regions of Queensland, respectively. In contrast, a weaker correlation between the daily new cases with RT-qPCR Ct value targets N1 or N2 (|r| ranging from 0.52∼0.23, Figure 4-d,e) was observed. These results suggest the sequencing method provides a more robust and consistent estimate of the prevalence trend compared to RT-qPCR across different regions.

### 3.4 Early detection and transition patterns of emerging VOCs

The emergence of SARS-CoV-2 variants with a growth advantage could lead to a greater infectivity or transmissibility and antibody escape, thus likely increasing the incidence rate and supplanting previous circulating variants.^3^ It is thought that public health measures effectively suppressed the local spread of these emerging variants in 2020/2021, evidenced by the fact that the low number of confirmed cases and the low RNA concentration in wastewater. For example, Brisbane entered three days of lockdown in response to the circulating Alpha variant (B.1.1.7, table S1) in January and late June 2021. Southeast Queensland implemented a 3-day lockdown and further extended it for an additional 5 days following the outbreak of the Delta variant (B.1.617) in August 2021. Correspondingly, these emerging variants were confined to a small cluster due to the timely implementation of measures. However, after the Queensland border reopened, the Omicron and its sublineages (BA*) transmitted rapidly and became dominant in Queensland (Figure S3).

In response to the limitation of extensive clinical surveillance, we explored the potential of sentinel sites wastewater sequencing as an alternative strategy to estimate the prevalence of circulating variants at the state level. To accomplish this, we compared the variant dynamics obtained in wastewater samples with that obtained in state-level clinical samples over the study period. Notably, variants detected in wastewater samples showed the main circulating variants were Omicron and its sublineages, with other co-occurring variants (e.g. Gamma and Delta) present in low abundance after the border reopened (Figure 5-a). The prevalence of the BA.2 variant replaced the BA.1.17 variant in mid-March 2022, and then later in July the BA.5* became dominant (Figure 5-a). This finding is consistent with the variant prevalence trend shown by clinical sequencing data (Figure 5-b). For example, clinical data showed that BA.2 replaced the BA.1.17 variant as the predominant circulating variant in Queensland in mid-March (Figure 5-b). More importantly, the detection of new variants in wastewater samples preceded their identification in clinical settings (Table S2). For instance, the parent Omicron variant (B.1.1.529) was detected in wastewater in February, 2022, but it was reported from clinical sequencing in March, 2022. Further, signals of the BA.2.75 variant were found in wastewater in April, 2022, preceding its local spread which was reported clinically in July, 2022. Moreover, the BA.3* variant was identified in wastewater but not in clinical sequencing, revealing the potential cryptic transmission of these variants in Queensland. Similarly, BQ.1 was detected in wastewater in late June, while it was not reported clinically until mid-September. It should be noted that we also observed the BA.2 variant was present in wastewater in early February 2022 but it had been observed in mid-January 2022 in confirmed clinical cases (Figure 5-a, b).

**Figure 5.**
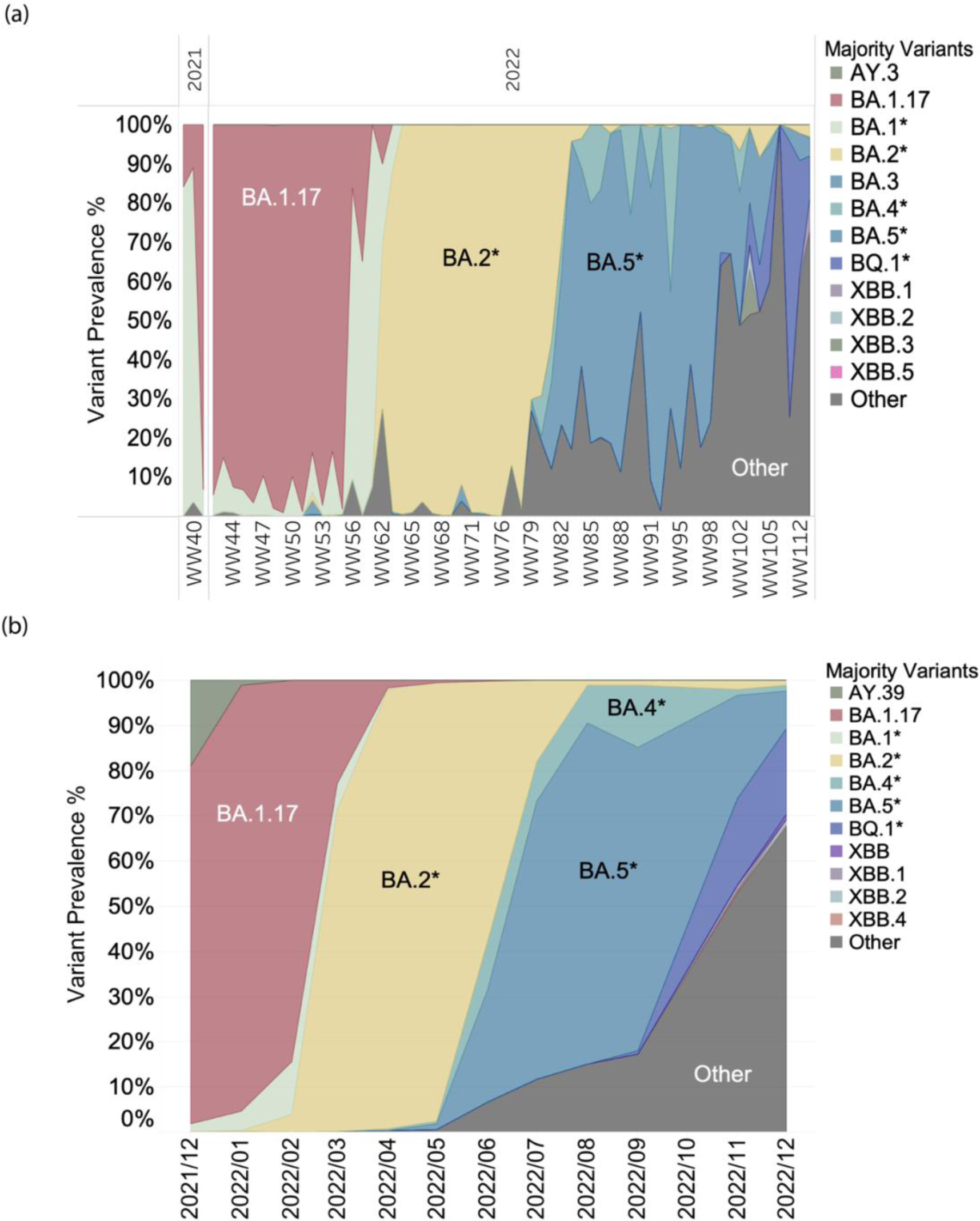
Prevalence of the SARS-CoV-2 variants identified in wastewater samples (a, n=64 wastewater-derived genomes with 10X coverage >20) and clinical samples (b, n=4878 GISAID sourced complete clinical genomes) during the experimental period expressed as a percentage. The different variants detected in wastewater are shown in different colours. The details of wastewater and clinical epidemiological data can be found in supplementary table S2.

### 3.5 SARS-CoV-2 genetic diversity in wastewater

The identification of SARS-CoV-2 variants relies heavily on the single nucleotide variants (SNVs) associated with each variant, thus genetic information is vital for monitoring the emergence of new variants.^1^ To determine the potential of wastewater sequencing in detecting the evolution of the virus, we calculated the genetic diversity in both wastewater and clinical samples by mapping the sequencing data against the reference genome (MN908947.3). To sum up, the mutation analysis of 65 wastewater samples and 4878 clinical genomes yielded 3473 and 17017 mutations, respectively, with a majority of mutations occurring in the S gene and ORF1ab gene (Figure 6-a, b). Genetic diversity metrics of richness and Shannon Entropy suggest the genetic diversity of SARS-CoV-2 in the clinical samples is greater than that in the wastewater samples (Figure 6-c). Figure 6-d depicts the 1469 mutations identified in the spike protein within wastewater samples. Among these mutations, 1155 are present in the S1 gene of the spike protein, which normally poses a high risk of instigating the emergence of novel variants (Figure S4). Notably, despite the higher genetic diversity observed in clinical samples, our analysis identified 28 unique spike protein mutations in wastewater samples, whereas they were absent in the clinical counterparts (Table S3).

**Figure 6.**
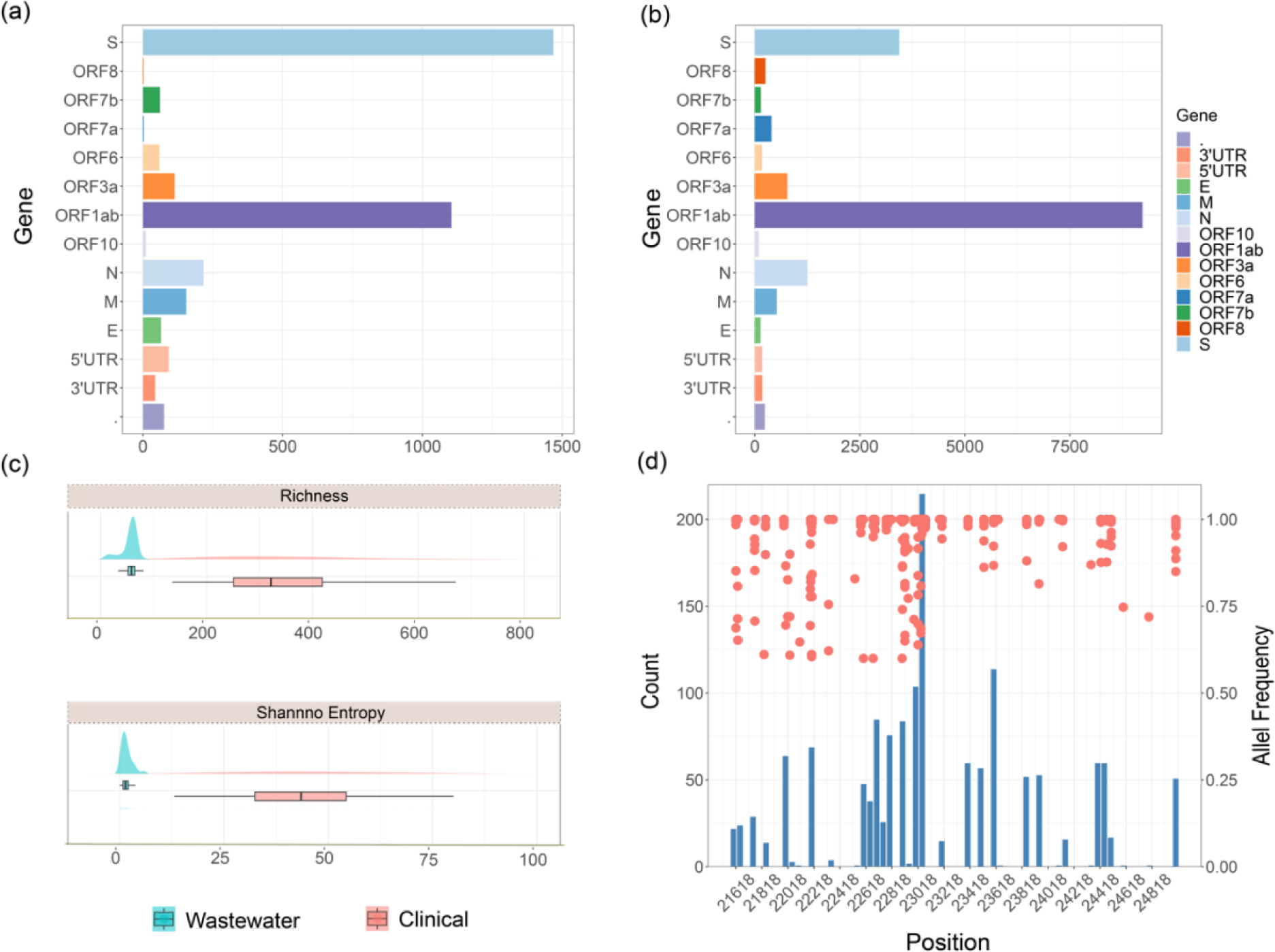
Summary of genetic variations in SARS-CoV-2 genes in wastewater and clinical samples. Count of mutations that occur on the different SARS-CoV-2 genes in wastewater samples (a) and clinical samples (b). Genetic diversity of SARS-CoV-2 in wastewater and clinical samples (c), represented by richness (top) and Shannon Entropy (bottom). Mutations detected in the Spike protein of SARS-CoV-2 in wastewater samples are shown in (d). The histogram (bin width = 50 bp) is the count of the number of each mutation that occurs on the genome position that is aligned with the left y-axis, while the scatter plot represents the allele frequency of each mutation in the corresponding position aligned with the right y-axis.

## 4. Discussion

This study highlights the capability of wastewater sequencing to accurately reflect the disease prevalence trend and the potential to provide early information on the circulating variant dynamics. These findings further support the WBE setting as an alternative surveillance program providing long-term, large-scale, and sustainable monitoring of SARS-CoV-2 across a wide geographic area. Moreover, the implementation of WBE as a surveillance tool holds significant promise in improving our preparedness and response strategies for future outbreaks and pandemics beyond SARS-CoV-2.

Although the PCR-based methods can provide timely and cost-effective estimation of disease prevalence, it is incapable of targeting emerging variants and providing genetic information to track virus evolution.^12,15,34–36^ In contrast, the ATOPlex sequencing method not only enables accurate quantification of the RNA concentration in wastewater but is also capable of identifying the co-occurrence of different VOCs, thus demonstrating wastewater as a valuable tool to monitor disease trends and variant dynamics in the community. This study found a stronger Pearson’s correlation between ATOPlex sequencing and number of daily new cases compared to the PCR-based method, particularly in the LG region (The correlation coefficients (|r|) for ATOPlex, N1, and N2 in relation to the daily new cases were found to be 0.66, 0.38, and 0.23, respectively. (Figure 4)). The higher dilution status in the SC region may have resulted in a decreased RNA signal, thus increasing the likelihood of RT-qPCR missing the target.^12^ In addition, an increasing disparity between the estimated viral concentrations in wastewater and the daily new cases reported in clinics has been observed. This can partially be explained by the gradual easing of public health measures, especially the widespread use of rapid antigen test (RAT) kits, which has led to a significant decline in the PCR-confirmed cases reported to clinics. Specifically, from mid-January to mid-February 2022, the PCR test number administered per 1,000 individuals per week in Queensland dropped from over 50 to approximately 15, which may account for the decrease in PCR-confirmed cases from 1,334.8 to 258.6 per 100,000 population.^37^ However, the correlation coefficient (|r|) between RNA concentration in wastewater and clinical parameters (e.g. daily or active total cases) varied in different studies, ranging from 0.14 to 1.^33^ This might be attributed to various evaluation conditions, such as environmental factors, sampling strategies and epidemiological factors.^33^ In addition, the Pearson correlation coefficient may be biased due to increased biases in the clinical data. Even though ATOPlex sequencing exhibits a higher |r| compared to the PCR method, further research is required to assess the potential impact of various factors on the strength of the correlation coefficient. Additionally, there is a need to optimize the prediction model to better understand the disease prevalence in the community.

Moreover, with millions of people still getting infected and thousands of deaths globally to date, ongoing investigations into the impact of COVID-19 variants and intervention policies (e.g., booster vaccination), are crucial due to the continuously changing circumstances. This longitudinal study demonstrates the potential of wastewater sequencing to track the transition of different variants, which aligns with clinical epidemiological reports in Queensland, Australia (Figure 5-a, b). Additionally, monitoring mutations in the SARS-CoV-2 genome is vital for informing health surveillance and response strategies, particularly in identifying and monitoring emerging variants.^25^ WBE as a means of monitoring mutations occurring in the SARS-CoV-2 genome is practical where large-scale clinical sequencing is unfeasible. Similar to clinical sequencing, the majority of mutations detected in wastewater were found in the receptor-binding domain (RBD) region of the spike protein, which has been identified as a key factor in the emergence of new variants (Figure 6-a,d).^38,39^ For example, the L452R mutation enhances the immune escape fitness and virulence of the Delta variant, while its absence in the Omicron variant reduces its pathogenicity.^38^ Furthermore, unique mutations were observed in multiple wastewater samples, revealing the hidden cryptic circulating variants in the community. These findings require careful interpretation to better understand the characteristics of SARS-CoV-2.

While obtaining high-quality genomes from wastewater samples is still challenging due to factors such as RNA degradation or low concentration, ATOPlex sequencing can recover complete or partial genomes from the wastewater samples, thus providing valuable retrospective epidemiological insights. However, certain regions with low sequencing depth were observed in the recently collected wastewater samples (Figure S2), underscoring the necessity to optimize the ATOPlex primer set used for amplicon sequencing to enable the investigation of emerging variants.^25^ Additionally, while retrospective analysis demonstrated the feasibility of early detection of emerging variants, it was observed that the genetic diversity obtained from regional wastewater was lower compared to the comprehensive clinical sequencing conducted across the entire state. This finding highlights the significance of selecting sampling sentinel sites with precision, particularly in areas characterized by high traffic or population density, as this is essential to ensure a comprehensive understanding of the circulating variants in the entire state. Future studies should focus on identifying appropriate sampling sites and establishing long-term routine wastewater sequencing monitoring, aiding effective public health decision-making and outbreak prevention.

## 5. Conclusions

This study aims to investigate the performance of using sentinel wastewater surveillance to track the variant dynamics at the state level. In this regard, we established a high-throughput, multiplex, amplicon-based sequencing technology, namely ATOPlex, to discern multiple variants from a longitudinal wastewater surveillance program in Queensland, Australia. The key conclusions are as follows:

- Comparison with a qPCR-based method showed that ATOPlex sequencing provides a more reliable snapshot of disease prevalence in a community. With the decrease in clinical tests, this method can assist public health sectors in comprehending the prevailing trends.
- Wastewater sequencing can track the variant dynamics over time and the results are comparable to the clinical sequencing results. It also has the potential to detect new variants before their proliferation, thus supporting the prevention of pandemics.
- Mutations can be identified in the recovered genome showing the ability to track virus evolution on a large scale.

These findings collectively suggest that sequencing-based WBE is a cost-effective and timely alternative strategy for monitoring diseases within a community.

## Acknowledgement

JG would like to acknowledge the Australian Research Council for funding support through Discovery Project (DP220101526). GN would like to acknowledge support from the Advance Queensland Industry Research Fellowship (RM2019002600) and an Early Career Postdoctoral Fellowship (ECPF23-8566329039) at the Faculty of Medicine, Nursing and Health Sciences of Monash University. We thank MGI Australia for sequencing support. The authors thank City of Gold Coast, Urban Utilities and Logan Water for their help in collecting samples. We thank Queensland Health for their help with data retrieval.

## Notes

The authors declare no competing financial interest.

