## Supplementary material for "Wastewater Tiling Amplicon Sequencing Reveals Longitudinal Dynamics of SARS-CoV-2 Variants Prevalence in the Community": Table S1, Figure S1, Figure S2, Figure S3, Figure S4

Figure S1: Wastewater sampling sites and positive control preparation.

Figure S2: Sequencing Depth of the wastewater samples.

Figure S3: Prevalence of the SARS-CoV-2 variants identified by clinical sequencing

Figure S4: Visualization of the S gene (MN908947.3:21,563~25,384) and the function of each structure.

Table S1: Key timeline of COVID-19-related events and public health response activities announced by the Queensland government.

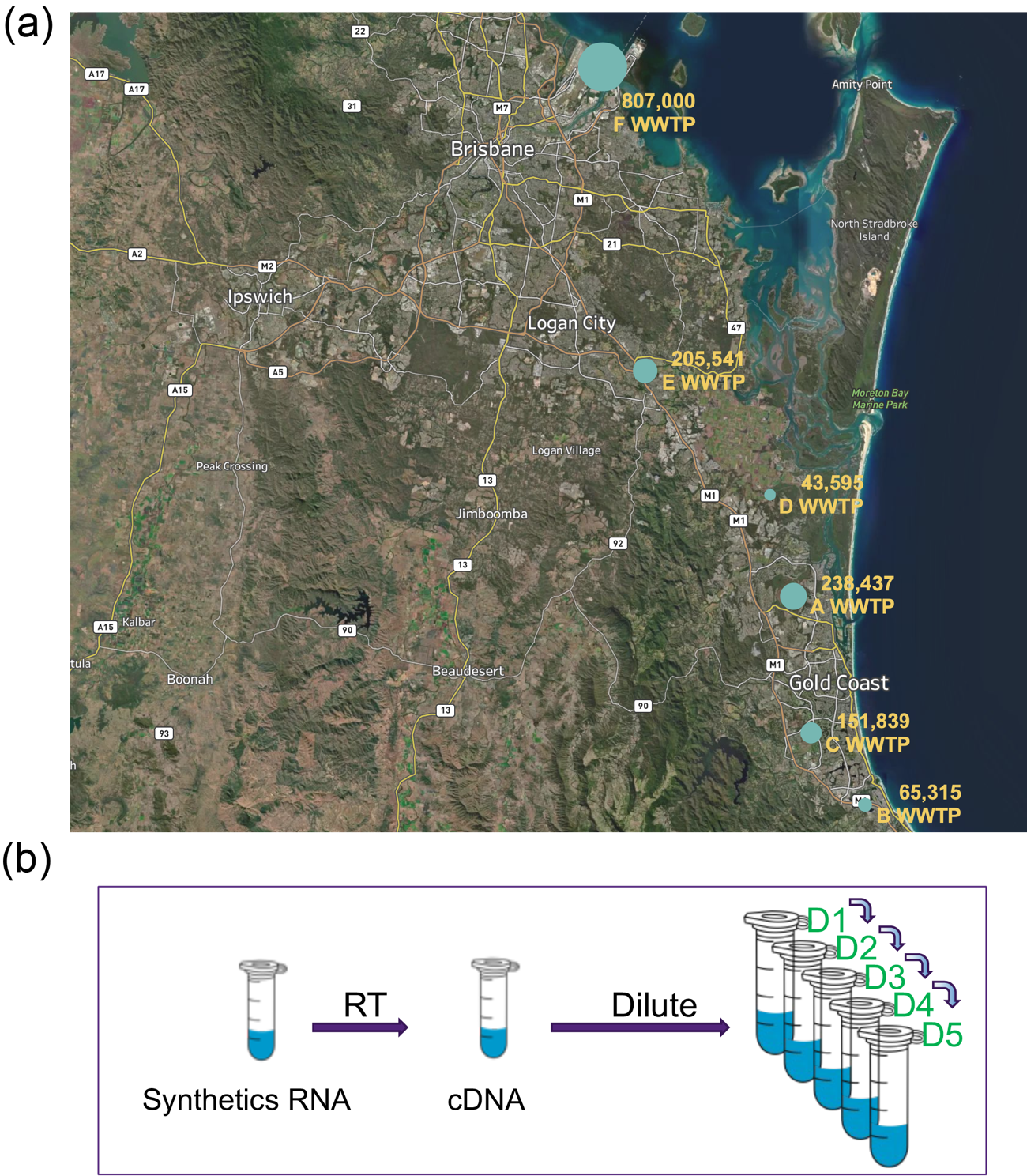

Figure S1. Wastewater sampling sites and positive control preparation. (a) Map of the sampling WWTPs. The dots on the map are the locations of the WWTPs and the number labelled below is the population served by the WWTP. (b) Positive control dilution series for sequencing.

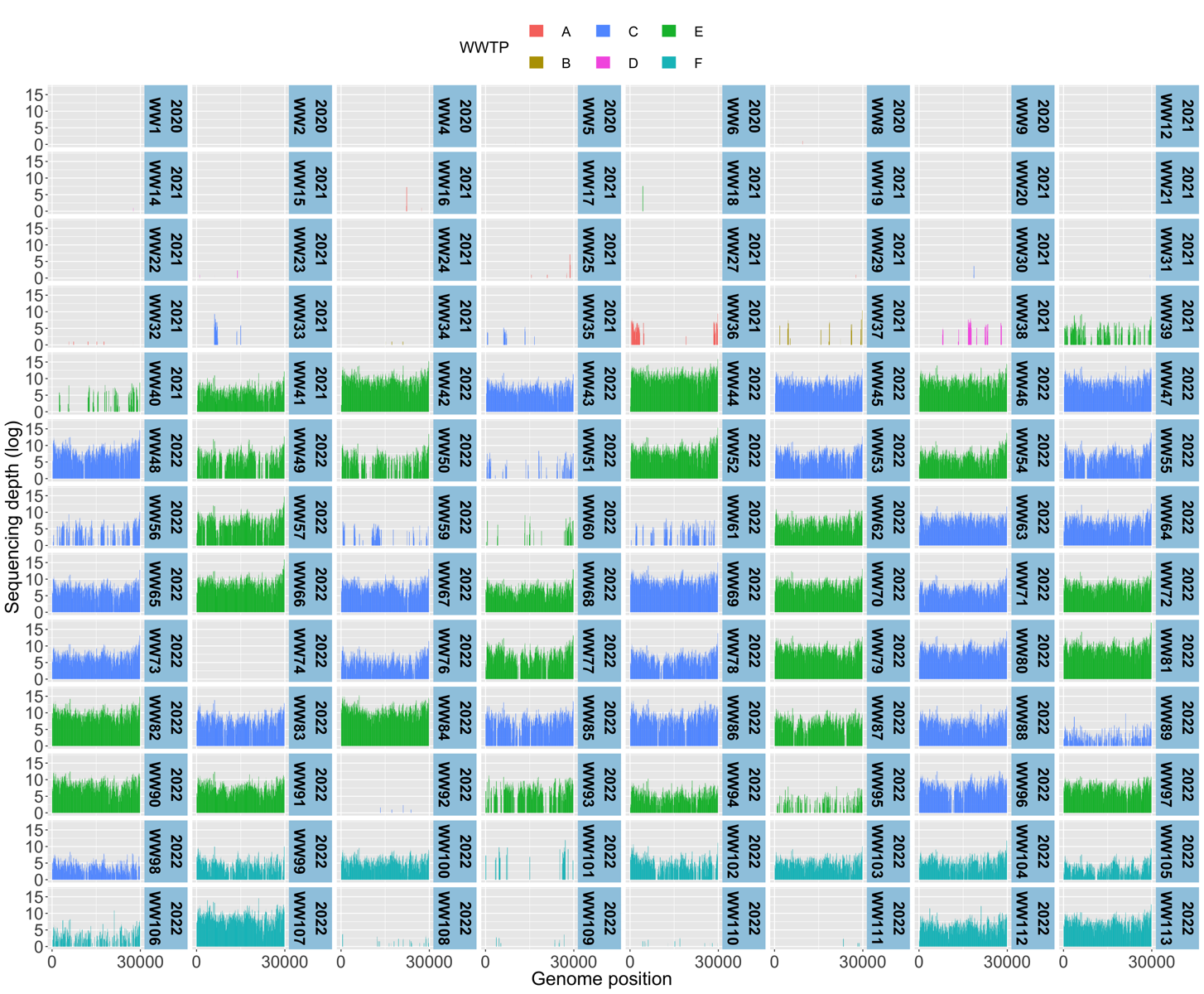

Figure S2 Sequencing Depth of the wastewater samples. The x-axis represents the nucleotide positions in respect to the reference genome (MN908947.3) and the y-axis represents the sequencing depth at each position.

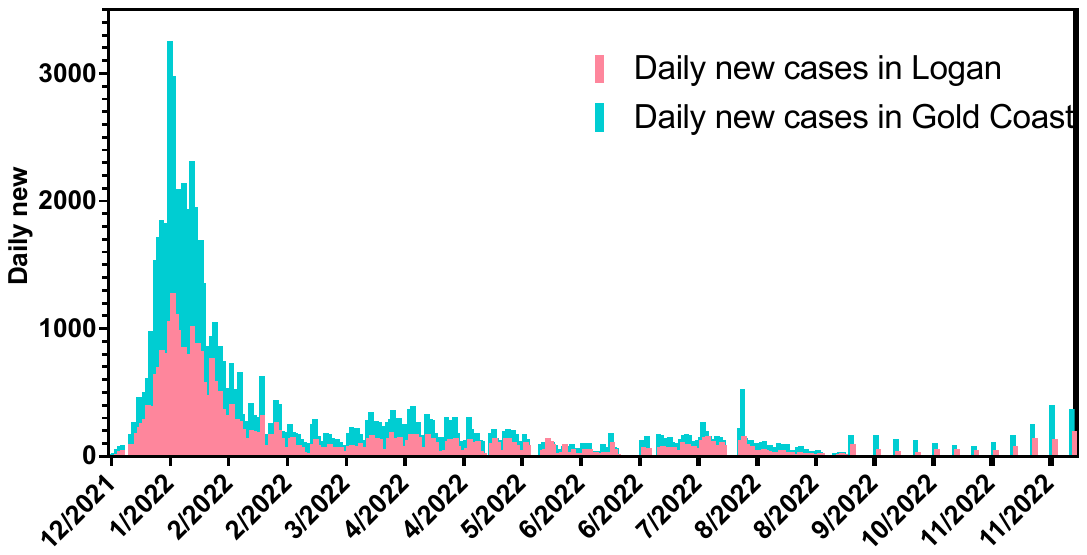

Figure S3 Prevalence of the SARS-CoV-2 variants identified by clinical sequencing. The dominant circulating variants in each epidemiology period are labelled.

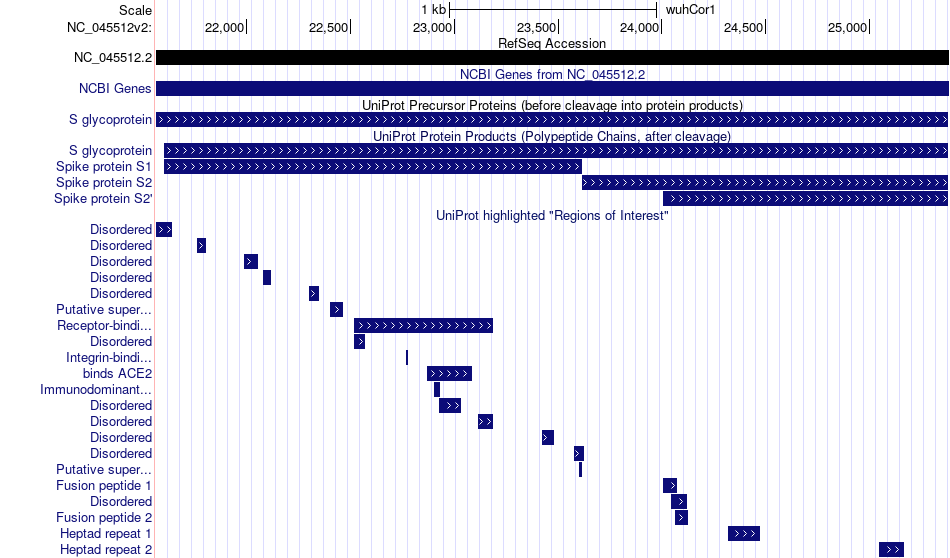

Figure S4 Visualization of the S gene (MN908947.3:21,563~25,384) and the function of each structure.^1^

Table 1 Key timeline of COVID-19-related events and public health response activities announced by the Queensland government.

| **Date** | **Event** | **Note** |
| --- | --- | --- |
| 1/20 | WHO declare Public health emergency of international concern. |  |
| 2/20 | Australia implemented the travel restrictions of China. |  |
| 2/20 | WHO announced the name for the new coronavirus disease: COVID-19. |  |
| 3/20 | WHO characteristic COVID -19 as pandemic. |  |
| 3/20 | Travel bans on foreign nationals entering Australia. |  |
| 3/20 | QLD border close for non-essential traveller. | Broder close |
| 3/20 | Social distance applied in QLD. |  |
| 3/20 | Australian citizens and Australian permanent residents are restricted from travelling overseas. |  |
| 3/20 | All people entering Australia are required to undertake a mandatory 14-day quarantine at designated facilities (e.g., hotels) in their port of arrival. |  |
| 3/20 | Both indoor and outdoor public gatherings limited to two persons only. |  |
| 5/20 | Gathering restrictions were eased. |  |
| 7/20 | QLD border reopened expect to Victoria. | Broder open |
| 7/20 | Gathering restrictions implemented. |  |
| 8/20 | QLD enhanced border measures. | Broder restriction. |
| 9/20 | Gathering restrictions implemented. |  |
| 9/20 | Gathering restrictions eased. |  |
| 10/20 | Gathering restrictions further eased. Queensland border zone no longer exists, New South Wales border zone extended. | Broder restriction was eased. |
| 11/20 | Restrictions were eased for NSW. |  |
| 12/20 | Broder close to NSW. | Broder close. |
| 1/21 | Greater Brisbane enters a three-day lock down. Face masks are mandatory. | Lockdown to 11/1/21; B.1.1.7 (Alpha) |
| 2/21 | Queensland closed the border to Victoria. | Broder close |
| 2/21 | Queensland opened the border to Victoria. | Broder open |
| 3/21 | Brisbane enters three days lock down. Queensland entered a period of higher level restrictions for Greater Brisbane. | Lockdown to 01/04/2021; Unknown community transmission. |
| 6/21 | Queensland announced all travellers from anywhere in Australia or New Zealand must complete a Queensland Travel Declaration. |  |
| 6/21 | Queensland mandated the Check in QLD app. |  |
| 6/21 | QLD announced a 3-day lockdown and extend 24h. | Lockdown to 03/07/2021; Alpha outbreak. |
| 8/21 | SEQ lockdown. | Lockdown to 08/08/2021; Delta outbreak. |
| 9/21 | Restrictions increased. |  |
| 12/21 | Qld borders re-open to domestic. | Broder reopen to all states; vaccine rate over 92.5% |
| 1/22 | Masks are still mandatory. |  |
| 10/22 | No Public Health Directions in effect. |  |
